# Improving esophageal protection during AF ablation: The IMPACT study

**DOI:** 10.1101/2020.01.30.20019158

**Authors:** Lisa WM Leung, Abhay Bajpai, Zia Zuberi, Anthony Li, Mark Norman, Riyaz Kaba, Zaki Akhtar, Banu Evranos, Hanney Gonna, Idris Harding, Manav Sohal, Nawaf Al-Subaie, John Louis-Auguste, Jamal Hayat, Mark M Gallagher

## Abstract

**Background:** Thermal injury to the esophagus is a known complication of ablation for atrial fibrillation (AF) and accounts for most procedure-related mortality. Thermal protection of the esophageal lumen by infusing cold liquid can reduce thermal injury to a limited extent. A method to control the local luminal esophageal temperature is investigated by this study.

**Objective:** To investigate the ability of a powerful temperature control device to protect the esophagus from ablation-related thermal injury.

**Methods:** A single-center, prospective, double-blinded randomized controlled trial was used to investigate the ability of the ensoETM device to protect the esophagus from thermal injury. This method was compared in a 1:1 randomization to a control group of standard practice utilizing a single-point temperature probe. In the study group, the device was used to keep the luminal temperature at 4°C during radiofrequency (RF) ablation for AF. Endoscopic examination was performed at 7 days post-ablation and esophageal injury was scored. The patient and the endoscopist were blinded to the randomization.

**Results:** We recruited 188 patients, of whom 120 underwent endoscopy. Thermal injury to the mucosa was significantly more common in the control group than in those receiving esophageal protection (12/60 versus 2/60; P=0.008), with a trend toward reduction in gastroparesis (6/60 Vs 2/60, p=0.27). There was no difference between groups in RF duration, force, power and combined ablation index (P value range= 0.2-0.9). Procedure duration and fluoroscopy duration were similar (P=0.97, P=0.91 respectively).

**Conclusion:** Thermal protection of the esophageal lumen significantly reduces ablation-related thermal injury compared to standard care. This method of esophageal protection is safe and does not compromise the efficacy of the ablation procedure.

**CONDENSED ABSTRACT:** Thermal injury to the esophagus causes most ablation-related deaths. We investigated the ability of a powerful method of esophageal temperature control to protect from thermal lesions during ablation, using a double-blinded randomized clinical trial to compare this to standard care. Patients randomised to receive thermal protection experienced significantly fewer lesions to the esophageal mucosa and a trend towards reduction in gastroparesis. Procedure efficacy and efficiency were not compromised.

## INTRODUCTION

Although its incidence is less than one per thousand cases, atrio-esophageal fistula (AEF) is the most common lethal complication of ablation performed for left atrial fibrillation (AF) or left atrial tachycardia (LAT), accounting for a majority of fatal complications from AF ablation and therefore a majority of all ablation-related mortality^1^. Both radiofrequency (RF) ablation and cryoablation have been associated with this complication^1,2^. Mild degrees of thermal injury to the esophagus can be seen on endoscopy after ablation in 10-30% of patients^3^, and the incidence of these mild lesions correlate with the risk of fistula. Symptomatic alterations to gastric motility from thermal injury to the peri-esophageal neural plexus occur between 5% and 74% of cases^4,5^. Many strategies have been proposed to lower the risk to the esophagus, but none has shown consistent evidence of effectiveness.

The ensoETM device (Attune Medical, Chicago IL) is routinely used to control body temperature in patients who are prone to hypothermia or hyperpyrexia in an intensive care setting, or whose body temperature must be lowered to protect an injured brain^6,7^. As it does so by warming or cooling the lumen of the esophagus and stomach, we hypothesised that it might protect the esophagus from localized thermal injury by controlling the local temperature.

## METHODS

### Trial design

The IMPACT study is a single-centre prospective double-blind randomized controlled trial that has been approved by the London-Stanmore Research Ethics Committee (IRAS ID 253844, NIHR CPMS ID 40619) and registered on ClinicalTrials.gov NCT 03819946.

### Study population

All patients attending or already listed for radiofrequency ablation for AF or LAT under general anaesthesia by participating electrophysiologists at our centre were screened. Patients who were unable or unwilling to undergo post-procedure endoscopy were excluded. Participants were randomly assigned in a 1:1 design to either receive thermal protection with the ensoETM device or standard care consisting of the use of a single sensor temperature probe. Patients were blinded to the treatment assignment **(figure 1)**.

**Figure 1:**
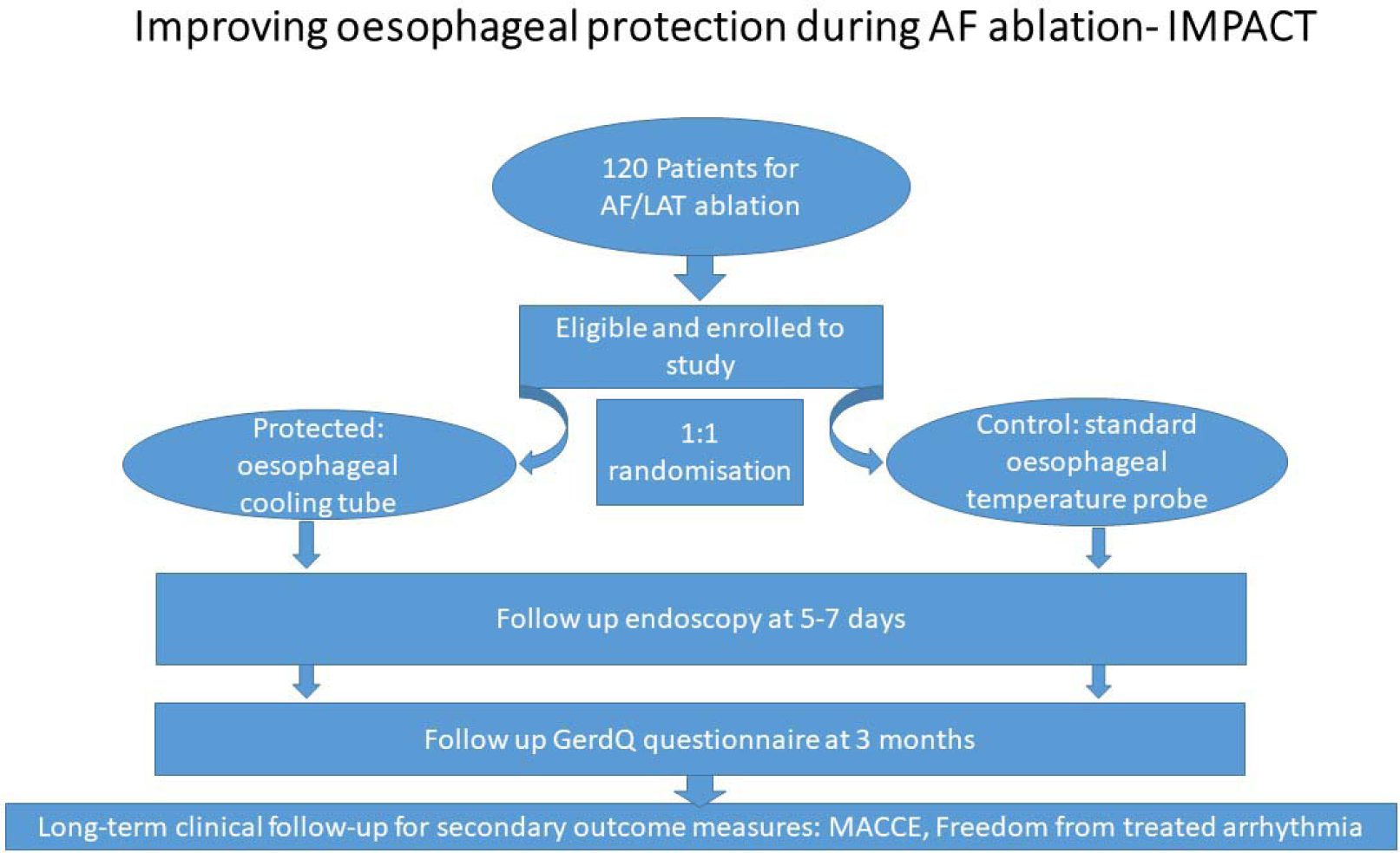
flowchart of the trial design and follow up.

### The ensoETM device

The ensoETM device is a silicone tubing (dimensions: 75cm in length, 1.2cm outer diameter; **figure 2**) that allows distilled water to flow in and out of the tube lumen in a closed loop manner; no foreign body or water enters the gastrointestinal tract of the patient. There is an additional inner lumen that can be used for gastric aspiration similar to a standard nasogastric (NG) tube. The non-patient end of the device is connected to a Blanketrol III mobile console (Gentherm Medical, Cincinnati, OH) and water is pumped from the console into the device. The water and therefore ensoETM device is set to the temperature required via the console. The water volume in the tubing is only 55ml and exerts a maximum pressure of 103kPa.

**Figure 2:**
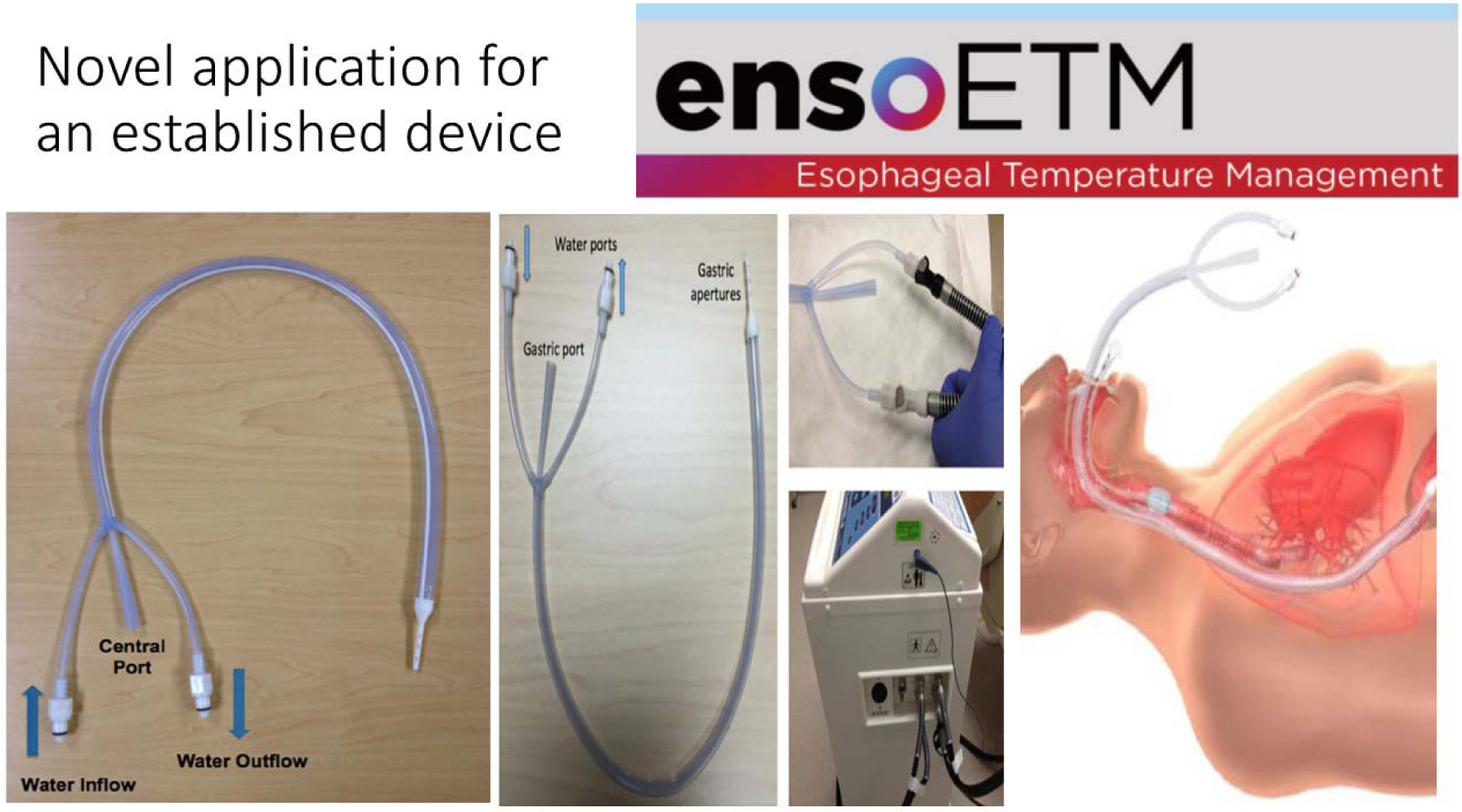
ensoETM device and Blanketrol III mobile console used to deliver controlled active thermal protection of the esophagus during AF ablation.

### Protected Group

Patients assigned to receive esophageal protection underwent preparation for ablation in the standard fashion with trans-esophageal echocardiography performed as soon as anaesthesia was induced. After using trans-esophageal echocardiography to guide transseptal puncture, the probe was withdrawn and an ensoETM probe was introduced in its place, connected to a Blanketrol III, mobile console. The position was confirmed radiographically. Before beginning ablation on the posterior part of the left atrium, the probe was set to cooling mode at 4°C for at least 10 minutes. Cooling continued until ablation was complete. External body temperature was recorded throughout via axillary approach or nasopharyngeal temperature probes.

### Control Group

A single-sensor temperature probe was placed in the esophagus by the attending anaesthetist and adjusted approximately to the site of ablation. Adjustment of the position of the probe during ablation was at the discretion of the operating electrophysiologist.

### Body temperature management in both groups

In all cases, standard anaesthetic methods were used, including endotracheal intubation, gas anaesthesia, whole body temperature monitoring and warming with a heated air-blanket (Bair Hugger™, 3M, St Paul, Mn, USA).

### Proton pump inhibitors

All participants in this study had proton pump inhibitors (PPI) prescribed post ablation for a period of 6-8 weeks. This is standard practice at our center.

### Method of RF ablation

Catheter ablation was performed using irrigated contact force sensing catheters (STSF or Qdot Micro, Biosense Webster, Johnson and Johnson, Diamond Bar, CA) with a 3D mapping system (Carto® version 7, Biosense Webster) with Ablation Index (AI) technology. Lesions in the anterior part of the left atrium were created at 40W with an AI target of 450-500; posterior lesions were at 30W with an AI target of 350-400.

### Endoscopy

All patients were invited to attend for endoscopy at 7 days after ablation. Endoscopy was performed by one of two senior endoscopists following a standardised protocol **(figure 3)** with detailed inspection of the anterior wall of the mid-oesophagus. The patient and the endoscopist were blinded to the treatment assignment of the patient. Any lesions observed were categorised according to a scoring system devised for the study. The scoring system was especially designed for the purpose of the study to improve delineation and characterisation of iatrogenic lesions caused by ablation compared to existing scoring systems for ulcerative lesions caused by different pathophysiology. The stomach was inspected for evidence of dysmotility. Gastroparesis was defined as the presence of food material in the stomach after more than 8 hours of fasting.

**Figure 3:**
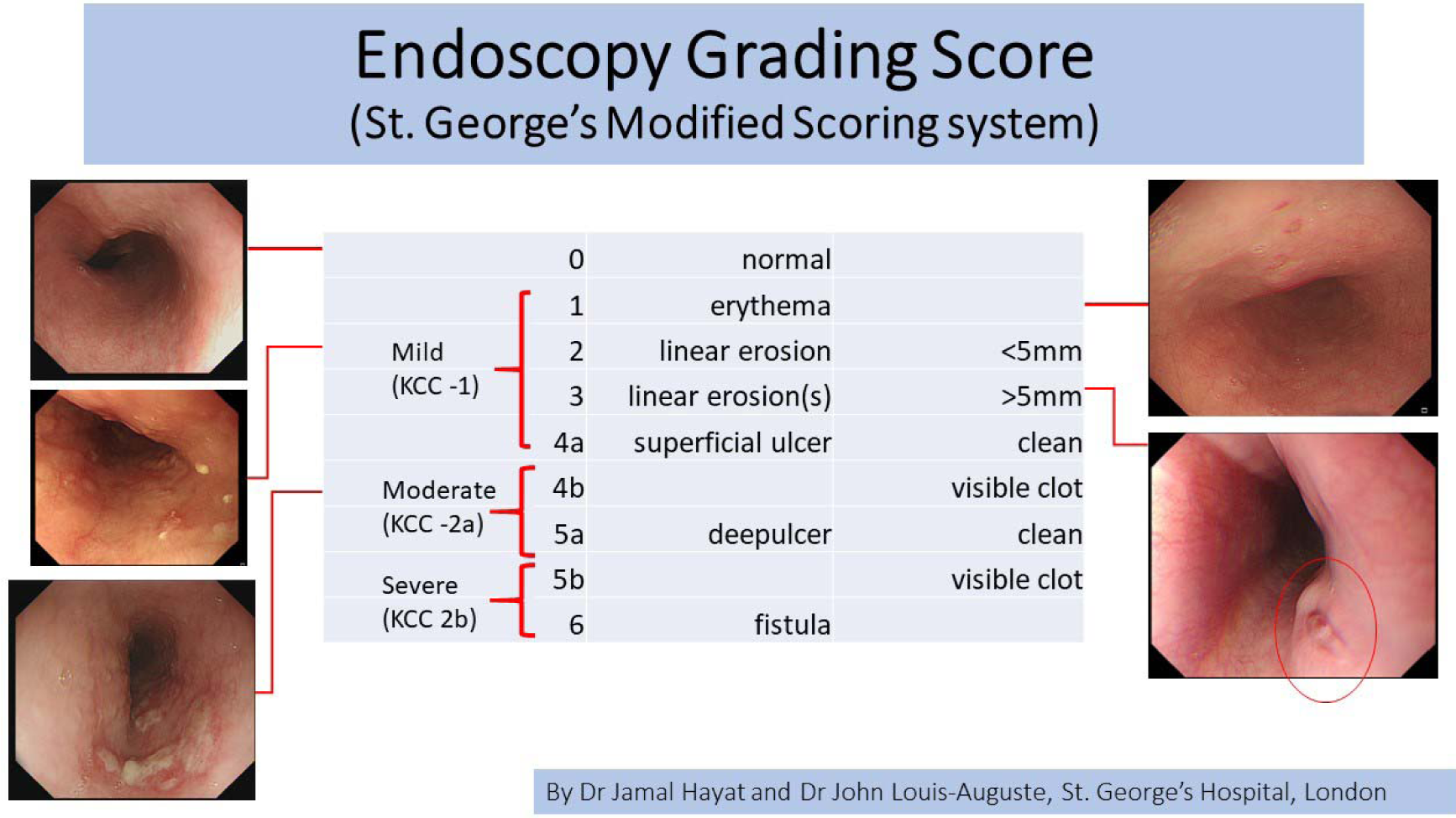
A St. George’s Modified Scoring System for endoscopically detected iatrogenic thermal mucosal lesions.

### Assessment of Symptoms

Patients underwent a structured assessment for upper GI symptoms according to the Gastro-Esophageal Reflux Disease (GERDQ) questionnaire at least 2 weeks after ablation. The Gastroparesis Cardinal Symptoms Index (GCSI) questionnaire was used to probe for typical features of gastric dysmotility and specific questions were used to quantify post-procedural chest pain. The follow up was conducted by members of the research study without access to the randomization of the participants.

### Arrhythmia Follow-Up

Patients were reviewed at 3 and 6 months post-ablation and underwent ambulatory ECG monitoring between these visits. Any occurrence of symptomatic or sustained AF or AT at >3 months post-ablation was considered to represent a failure of the procedure.

### Power Calculation

Based on a pre-study clinical estimate of 15% between the study and control groups, we calculated a sampling population size of 120 with endoscopic assessment to achieve a study power of 0.80 to answer the hypothesis.

### Endpoints

The primary endpoint of the study was incidence of endoscopically detected esophageal mucosal lesions and/or gastroparesis. The timing of endoscopic examination was 7 days. The main secondary endpoints included the presence of significant patient chest and gastroenterological symptoms as recorded systematically in the validated questionnaires GERD-Q and GCSI. Timing of questionnaire assessment was earliest at 2 weeks to >3 months post ablation. (graded 0-18; 0-45. 18 and 45 being the worst possible scores respectively). We also recorded the incidence of major adverse cardiovascular cerebrovascular events (MACCE): MI/CVA, all-cause mortality, vascular trauma needing surgery, cardiac tamponade, AEF, hospital acquired infection. Assessment occurred at 0, 3 and 6 months post-ablation. 12 month follow up will also be conducted.

The acute success of the procedure was defined as isolation of the pulmonary veins and proven block across any other line of lesions delivered; clinical success was also measured as freedom from the treated arrhythmia at >3 months after ablation. The duration of the procedure and duration of fluoroscopy was documented in all cases, as well as ablation delivery parameters including total RF ablation time, power, force, FTI and combined ablation index. Standard ECG monitoring devices were used for follow-up, including 12 lead ECG at all follow-up visits in all patients, implantable loop recorders (ILR) and CRM devices where available, ambulatory ECG of ≥24 hours in all other patients.

### Statistics

Analysis was performed with IBM SPSS statistical software (Version 22.0, IBM SPSS Statistics, NY, USA) using chi□square test and Fisher’s exact test as appropriate. Kaplan–Meier analyses with log□rank tests were used to calculate AF/study arrhythmia recurrence□free survival over time and compare recurrence rates across groups. Logistic regression analyses was used to identify predictors of specific lesion durability e.g. LAPW isolation. Cox regression analyses was be used to assess independent predictors of AF and/or other study arrhythmia recurrence after catheter ablation.

## RESULTS

Patients were recruited from 22nd Feb 2019-13th January 2020. More than 90% of patients screened for recruitment agreed to participate, but 36.2% of recruited patients later expressed unwillingness or inability to return for endoscopy. A total of 188 participants were recruited (93 protected, 95 control) of whom 120 (60 protected, 60 control) underwent endoscopic examination. All 188 participants underwent structured follow-up for post-ablation symptoms and clinical outcome as per protocol.

### Patient and procedure characteristics

The baseline characteristics of the groups were well matched **(table 1)**. The use of esophageal cooling was not associated with any difference in the procedure duration, nor was there any evidence that its use made the accomplishment of procedural endpoints more difficult. All veins were isolated in all subjects in both groups, and all veins remained isolated at the end of the procedure.

**Table 1:** Patient and procedure characteristics.

| Patient & Procedure Characteristics | PROTECTED (n=60) | CONTROL (n=60) |
| --- | --- | --- |
| Male | n=36(60%) | n=37 (61.7%) |
| Age (years) | 64.5 | 64.7 |
| LA diameter - anteroposterior (cm) | 4.1cm | 4.2cm |
| LV ejection fraction (Simpson's) | 50.50% | 52.30% |
| BMI (kg/m2) | 28.5 | 29.8 |
| Persistent, 1<sup>st</sup> time ablation | n=27 (45%) | n= 30 (50%) |
| PAF, 1<sup>st</sup> time ablation | n=24 (40%) | n=20 (33.4%) |
| Repeat left atrial ablation | n=8 (13.3%) | n=9 (15%) |
| LAT (left atrial tachycardia) | n=1 (1.7%) | n=1 (1.7%) |

Linear ablation lesion sets across the roof of the left atrium, the posterior wall of the left atrium and the posterior mitral isthmus were each attempted at similar proportions of the protected cases and in the control group; success at these sites was similar in both groups. The total number of lesions and the duration of RF delivery required to achieve procedural endpoints was similar in the control and the protected group, both among those who received PVI only and amongst those receiving more extensive lesion sets **(table 2 and table 3)**.

**Table 2:** Procedural metrics.

| Procedure Metrics | Protected (n=60) | Control (n=60) | p |
| --- | --- | --- | --- |
| Procedure Duration (minutes) | 185.8 +/- 47.3 | 186.6 +/- 47.7 SD | ns |
| Fluoroscopy Duration (minutes) | 10.9 +/- 7.3 | 11.1 +/- 7.4 | ns |
| Achievement of PVI | 60/60 | 60/60 | ns |
| Posterior wall isolation | 16/23 | 19/23 | ns |
| Acute Complications | 0 | 2 | ns |
| Hospital Stay > 1 night | 1 | 1 | ns |
| MACE – within 3 months | 0 | 0 | ns |
| MACE – within 6 months | 1 | 0 | ns |

**Table 3:** Left atrial posterior wall ablation treatment delivery between protected group and controls.

|  | Duration of RF (min) | Average Force (g) | Max Temperature (°C) | Max Power (W) | Baseline Impedance | Impedance Drop | FTI | RFIndex |
| --- | --- | --- | --- | --- | --- | --- | --- | --- |
| **Control (mean)** | 14.1 | 19.1 | 26.6 | 33.9 | 122.0 | 8.6 | 248 | 394 |
| **SD** | 5.5 | 9.8 | 1.9 | 5.3 | 14.2 | 4.9 | 111 | 67 |
| **Protected (mean)** | 14.5 | 17.8 | 27.2 | 34.1 | 130.0 | 7.9 | 220 | 384 |
| **SD** | 6.5 | 10.3 | 2.3 | 4.7 | 11.9 | 3.9 | 99 | 54 |
| ***P value*** | *0.9* | *0.8* | *0.6* | *0.9* | *0.2* | *0.7* | *0.6* | *0.7* |

### Primary endpoint analysis - Endoscopy findings

Endoscopy demonstrated a significant excess of thermal injuries in the control group compared to those protected by the ensoETM (12/60 vs 2/60 p=0.008; **figure 4**). Gastroparesis was present in 6/60 patients in the control group and 2/60 protected patients (p=0.27). There were no endoscopy findings consistent with TEE trauma or from the ensoETM device. Severe (grade 4 or more) lesions occurred in only one case in the protected group: A grade 4a lesion that related to a protocol breach. The patient exhibited a recurrence of conduction into the right pulmonary veins as the procedure was finishing, and the operator delivered additional lesions at a site on the posterior margin of the right inferior vein, unaware that the esophageal cooling had been turned off 20 minutes earlier. The event was discussed after the protocol breach was noticed, but the patient continued in the study as the intention had been to protect throughout the procedure. The endoscopist was unaware of the treatment assignment or the deviation from protocol.

**FIGURE 4.**
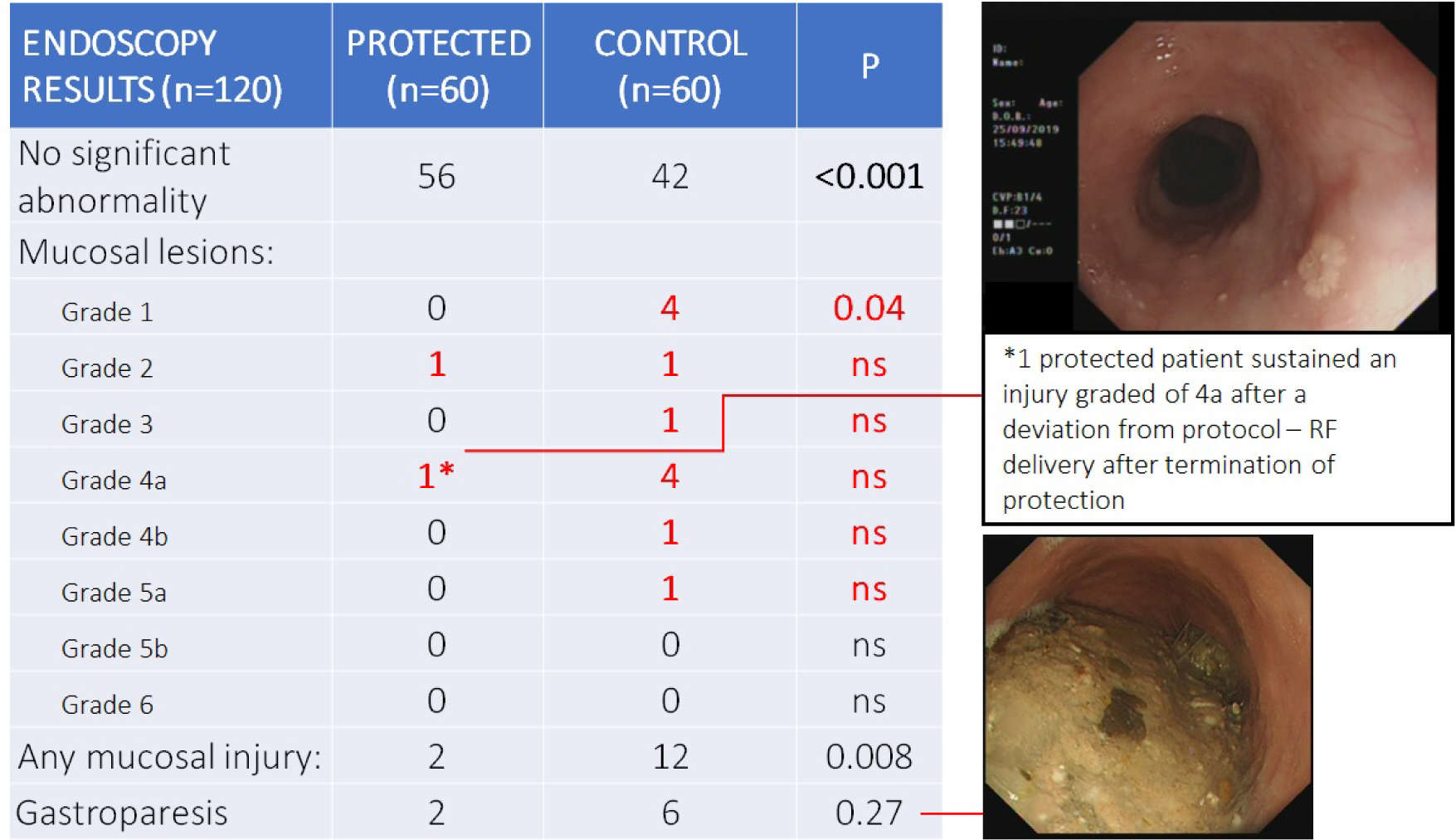
PRIMARY ENDPOINT: ENDOSCOPY FINDINGS.

### Secondary endpoint analyses - Post-ablation symptoms

Symptom scores for all gastrointestinal symptoms based on the GERD-Q and GCSI validated questionnaires were broken down point by point and the scores between the 2 groups were compared. In addition, chest pain symptoms post ablation patient experience was recorded on a scale of severity from 0-10 (10=most severe or worst experience).

### Clinical outcome

Recurrence of AF or AT outside the blanking period was equally common in the protected group versus the control group during follow up at 6 months. Long term follow up of the study subjects is ongoing.

### Major adverse cardiovascular cerebrovascular events (MACCE)

Time point of measurement of MACCE was at 0, 3 and 6 months post ablation. Acutely, there was no significant difference in MACCE rates in the protected and control groups.

There were 2 cases of acute vascular trauma needing intervention with thrombin injection in the control group and 1 case of additional hospital night stay due to post procedural bradycardia. There were 2 cases of extra hospital night stay in the protected group, of which one was planned pre-ablation due to previous adverse reaction to Heparin. The other case was due to a small pericardial effusion that was conservatively managed. There were no acute cases of tamponade, MI, CVA or death in either group. There were no cases of AEF recorded.

There was 1 case of late mortality in the protected group, a patient who died at between 3 and 6-months post ablation due to progression of pre-existing heart failure.

### Procedural workflow

Fluoroscopy and procedure duration were similar in both groups. Recovery time was also similar with no difference in hospital night stay between the 2 groups **(Table 2)**.

## DISCUSSION

This prospective double-blind randomised controlled trial demonstrates that endoscopically detected thermal injury to the esophagus is less frequent when the ensoETM device is used to control the local temperature of the esophageal lumen during RF ablation for AF. This is the first randomized clinical study to show superiority of this method of esophageal protection over standard care.

### Secondary endpoints

The analyses on the secondary endpoints show that the ensoETM device is safe, with no difference in MACCE rates or acute complications when compared to the control group cases. Endoscopy findings also did not show any evidence of esophageal trauma attributable to the ensoETM device or to trans-esophageal echo. Procedure work-flow and acute success were unaffected by the device and at the 6-month time point, there is no difference between the groups in the efficacy of the ablation.

The evaluation of new technologies can impair procedural workflow but despite the novelty of the ensoETM, we have demonstrated that esophageal cooling did not adversely impact procedure or fluoroscopy times.

Operators participating in the study could not have been blinded to the randomization. We were conscious of the risk that a perceived protection might foster recklessness by the operators, so all were exhorted to follow as closely as possible to their standard methods and lesion sets. The data collected on ablation power, contact force, time, impedance, FTI and combined Ablation Index confirm that they did so. This strict adherence to standard methodology may have contributed to the high level of esophageal protection observed. It should not be interpreted as a licence to abandon all restraint in ablating the region close to the esophagus.

### Thermal Injury to the Esophagus

Lesions of the esophageal mucosa are common after AF ablation. This is widely recognised for RF, but also well documented for those who receive cryoablation. Schemes of classification have been devised to reflect this gradation of prognostic importance. The commonly used Kansas City Classification^8^ has 3 levels of severity; our more refined classification of thermal lesions with its 8 grades of injury provides a high level of discrimination which we believe to be useful in demonstrating trends in lesion severity as well as incidence.

Our trial was unusual among studies of post-ablation esophageal injury in choosing a 7-day time point for endoscopy. Most thermal lesions are transient, consisting of erythema that resolves within days. Mechanical injury to the esophagus from echo probes, temperature probes or protective devices would be expected to follow a similar time-pattern. Moderately severe lesions appear soon after ablation but heal within weeks. Ulceration of the mucosa is known to be linked to progression towards AEF formation^9, 10^, a complication that typically presents clinically at between 2 and 4 weeks after ablation.

We chose 7 days as the ideal interval between ablation and endoscopy based on the available literature as the optimum for detecting significant lesions without being overwhelmed by trivial findings. A systematic review showed that with up to 47% incidence of all mucosal lesions at endoscopy performed at 24 hrs post procedure^3,11,^, differentiation between minor lesions and those worthy of attention was difficult; endoscopy performed at an interval of 7-14 days from the time of ablation gave far greater discrimination^9^. For this reason and the known variable course of severe mucosal lesions and AEF formation, the timing of endoscopic evaluation in our study was set to 7 days.

We recognise that even this approach with endoscopy assessment will still have limitations. There is still a risk of falsely declaring a case as ‘all clear’; previous reports show anomalous cases having no mucosal lesions detected at endoscopy 18 days post ablation but nonetheless progressing to AEF^5^. Despite this limitation, endoscopic examination is still our best diagnostic tool. Computerised Tomography has a poorer performance, with far greater numbers of false negative reports but is a recognised gold standard diagnostic test for emergent cases of suspected AEF^12^.

### Preventing Atrio-Esophageal Fistula

Strategies have been tried for the prevention of atrio-esophageal fistula including the administration of proton-pump inhibitors to facilitate oesophageal healing. This has intuitive appeal but no backing in trial evidence^3,13^. It is based on the assumption that gastric acid contributes to the injury that progresses to AEF formation; it is equally likely that bacterial action is the prime driver for this progression and acidity might be regarded as protective in its bactericidal effect.

Operator restraint in the power delivered to the left atrial posterior wall was the first strategy shown to reduce the risk of AEF. More recently, limitation in the use of contact force has been shown to help^14^. Unfortunately, this conservative methodology risks producing less effective lesion sets, hindering procedural success as well as contributing to less efficient workflow.

Mechanical deviation of the esophagus away from the point of ablation has been investigated extensively. The most common method involves manipulation of the trans-esophageal echo probe to deviate the esophagus^15^. Dedicated devices have also been studied Balloon Retractor^16^. These methods are imperfect and potentially harmful. A recent large multi-centre study showed that trans-esophageal echo probe insertion and manipulation is an important cause of mortality in general anaesthetic cases undergoing cardiothoracic surgery or cardiac procedures with an incidence of 1 in 3,000 and a mortality rate of 40% to those that sustained esophageal injury^17^. Mechanical deviation of the esophagus by any device could cause similar complications.

### Previous studies on esophageal cooling

Previous small studies have investigated the efficacy of esophageal cooling during ablation. Although individually inconclusive, taken together, these show clear evidence of protection, but the effect is small^18,19^. The small magnitude of the protection is unsurprising, given the limited heat extraction capability of the methods used. Esophageal cooling using the ensoETM device is simple and easily standardised; it is also a much more effective heat extractor than the methods investigated by previous studies.

### Gastric Motility

Previous post-ablation endoscopic studies were either focused on mucosal lesions or functional pathology^4,20^. Our trial protocol included a detailed assessment of gastric motility as well as the esophageal mucosa. The IMPACT study shows a non-significant excess of both endoscopically defined delay in gastric emptying and in symptoms related to this condition in the control group compared to patients receiving thermal protection.

## CONCLUSION

Active control of the esophageal luminal temperature by the ensoETM device can protect the esophagus from thermal injury during left atrial ablation for AF or AT and may also protect its associated neural plexus.

## LIMITATIONS

We have not proved that the use of the ensoETM device eliminates the possibility of atrio-esophageal fistula formation. A large international multi-centre randomized clinical trial is required to offer insight towards the level of esophageal protection offered by utility of this device compared to current control methods. In addition, a sense of security can foster recklessness; the operator performing AF ablation must consciously guard against this phenomenon.

## Data Availability

The study data is available to all study investigators at the Institution. It has not been published by any journal. Preliminary results have been presented as abstracts at the AHA 2019 Featured EP Science session and at the AF Symposium 2020, Washington DC.

https://www.ahajournals.org/doi/10.1161/CIR.0000000000000742

## ETHICAL APPROVAL

ETHICALLY APPROVED BY THE LONDON-STANMORE RESEARCH ETHICS COMMITTEE (IRAS ID 253844).

## REFERENCES

1. Kapur S, Barbhaiya C, Deneke T, Michaud GF. Esophageal Injury and Atrioesophageal Fistula Caused by Ablation for Atrial Fibrillation. Circulation; 2017; 136(13): 1247–1255.

2. Miyazaki S, Nakamura H, Taniguchi H, Takagi T, Iwasawa J, Watanabe T et al. Esophagus-Related Complications During Second-Generation Cryoballoon Ablation-Insight from Simultaneous Esophageal Temperature Monitoring from 2 Esophageal Probes. J Cardiovasc Electrophysiol. 2016; 27(9):1038–44.

3. Schmidt M, Nolker G, Marschang H, Gutleben KJ, Schibgilla V, Rittger H et al. Incidence of oesophageal wall injury post-pulmonary vein antrum isolation for treatment of patients with atrial fibrillation. Europace. 2008;10(2):205–9.

4. Lakkireddy D, Reddy YM, Atkins D, Rajasingh J, Kanmanthareddy A, Olyaee M et al. Effect of atrial fibrillation ablation on gastric motility: the atrial fibrillation gut study. Circ Arrhythm Electrophysiol. 2015; 8(3): 531–6.

5. Park, SY, Camilleri M, Packer D, Monahan K. Upper Gastrointestinal Complications Following Ablation Therapy for Atrial Fibrillation. Neurogastroenterol Motil. 2017; 29(11):10.1111/nmo.13109

6. Hegazy AF, Lapierre DM, Butler R, Martin J, Althenayan E. The esophageal cooling device: A new temperature control tool in the intensivist’s arsenal. Heart and Lung. 2017; 46(3); 143–148.

7. Bhatti F, Naiman M, Tsarev A, Kulstad E. Esophageal Temperature Management in Patients Suffering from Traumatic Brain Injury. Therapeutic Hypothermia and Temperature Management. 2019; 9(4). https://doi.org/10.1089/ther.2018.0034

8. Yarlagadda B, Deneke T, Turagam M, Dar T, Paleti S, Parikh V et al. Temporal relationships between esophageal injury type and progression in patients undergoing atrial fibrillation catheter ablation. Heart Rhythm. 2019; 16(2):204–212.

9. Keshishian J, Young J, Hill E, Saloum Y, Brady PG. Esophageal Injury Following Radiofrequency Ablation for Atrial Fibrillation. Gastroenterol Hepatol. 20128(6):411–414.

10. Doll N, Borger MA, Fabricius A, Stephan S, Gummert J, Mohr FW et al. Esophageal perforation during left atrial radio-frequency ablation: is the risk too high? J Thorac Cardiovasc Surg. 2003;125:836–842.

11. Di Biase L, Saenz LC, Burkhardt DJ Vacca M, Elayi CS, Barrett CD et al. Esophageal capsule endoscopy after radiofrequency catheter ablation for atrial fibrillation: documented higher risk of luminal esophageal damage with general anesthesia as compared with conscious sedation. Circ Arrhythm Electrophysiol. 2009; 2(2):108–12.

12. Chavez P, Messerli FH, Dominguez AC, Aziz EF, Sichrovsky T, Garcia D et al. Atrioesophageal fistula following ablation procedures for atrial fibrillation: systematic review of case reports. Open Heart. 2015;;2:e000257. doi: 10.1136/openhrt-2015-000257

13. Zellerhoff S, Lenze F, Eckardt L. Prophylactic proton pump inhibition after atrial fibrillation ablation: is there any evidence? EP Europace. 2011; 13(9): 1219–1221.

14. Zhang X, Kuang X, Gao X, Xiang H, Wei F Liu T et al. RESCUE-AF in Patients Undergoing Atrial Fibrillation Ablation. Circ Arrhythm Electrophysiol. 2019; 12:e007044.

15. Herweg B, Johnson N, Postler G, Curtis AB, Barold SS and Iiercil A. Mechanical esophageal deflection during ablation of atrial fibrillation. Pacing Clin Electrophysiol. 2006; 29(9): 957–61.

16. Bhardwaj R, Naniwadekar A, Whang W, Mittnacht AJ, Palaniswamy C, Koruth JS et al. Esophageal Deviation During Atrial Fibrillation Ablation. JACC Clin Electrophysiol. 2018 4(8):1020–1030.

17. Ramalingam G, Choi SW, Agarwal S, Kunst G, Gill R, Fletcher SN et al. Complications related to peri-operative transoesophgeal echocardiography-a one-year prospective national audit by the Association of Cardiothoracic Anaesthesia and Critical Care. Anaesthesia. 2020; 75 (1): 21–26.

18. Leung LW, Gallagher MM, Santangeli P, Tschabrunn C, Guerra JM, Campos B et al. Esophageal cooling for protection during left atrial ablation: a systematic review and meta-analysis. J Interv Card Electrophysiol. 2019. https://doi.org/10.1007/s10840-019-00661-5

19. Tsuchiya T, Ashikaga K, Nakagawa S, Hayashida K, Kugimiya H. Atrial Fibrillation Ablation with Esophageal Cooling with a Cooled Water-Irrigated Intra-esophageal Balloon: A Pilot Study. J Card Electrophysiol. 2007; 18(2): 145–150.

20. Knopp H, Halm U, Lamberts R, Knigge I, Zachaus M, Sommer P et al. Incidental and ablation-induced findings during upper gastrointestinal endoscopy in patients after ablation of atrial fibrillation: a retrospective study of 425 patients. Heart Rhythm. 2014; 11(4):574–8.

